# On the Fidelity versus Privacy and Utility Trade-Off of Synthetic Patient Data

**DOI:** 10.1101/2024.12.06.24317239

**Authors:** Tim Adams, Colin Birkenbihl, Karen Otte, Hwei Geok Ng, Jonas Adrian Rieling, Anatol-Fiete Näher, Ulrich Sax, Fabian Prasser, Holger Fröhlich, Alzheimer’s Disease Neuroimaging Initiative

## Abstract

The use of synthetic data is a widely discussed and promising solution for privacy-preserving medical research. Synthetic data may however not always be privacy preserving and can vary greatly in terms of data fidelity and utility.

We systematically evaluate the trade-offs between privacy, fidelity, and utility across five synthetic data models and three patient-level datasets. We evaluate fidelity based on statistical similarity to the real data, utility on three machine learning use cases and privacy via membership inference, singling out, and attribute inference risks. Synthetic data without differential privacy (DP) maintained fidelity and utility without evident privacy breaches, whereas DP-enforced models significantly disrupted correlation structures. K-anonymity-based data sanitization, while preserving fidelity, introduced notable privacy risks. Our findings emphasize the need to advance methods that effectively balance privacy, fidelity, and utility in synthetic patient data generation.

**Highlights:**

- Differential Privacy (DP) had a detrimental effect on feature correlations
- Models that did not implement DP showed good fidelity compared to real data
- Non-DP synthetic models showed no strong evidence of privacy breaches
- k-anonymization produced high fidelity data but showed notable privacy risks

## 1. Introduction

With recent advances and a growing number of applications of machine learning (ML) and artificial intelligence (AI), the availability and sharing of health data is of utmost importance and could potentially generate huge value for medical research and healthcare In an effort to make health related data available for research, there exist numerous initiatives such as the Alzheimer’s Disease Neuroimaging Initiative (ADNI) [1] or the Cancer Genome Atlas (TCGA) [2]. These data initiatives have been shown to have a big impact on research and inspire a large amount of follow up publications [3], which can in turn significantly advance research in the corresponding field [4].

Even though data sharing has been shown to elevate research, working with health-related data and in particular patient-level data is a delicate topic due to its sensitivity and the resulting privacy concerns. Due to such concerns and the corresponding legal data protection requirements [5], sharing health data is often a cumbersome and slow process. This is especially the case when attempting to share data across different institutions, which typically requires complex legal agreements between organizations that may not have the same interests. In consequence, privacy concerns can aggravate the formation of fragmented data silos. While approaches such as Federated Learning (FL) [6] try to address these challenges by allowing model training across decentralized data sources without exchanging raw data, there still persist significant challenges [7]. In addition to organizational, legal and technical issues, implementation of ML algorithms within a FL environment is impossible without data scientists having the possibility to see sample data. While randomly permuted real data or data sampled uniform randomly from the theoretical ranges of each individual variable can at least provide some insights into the principle structure of the real data, “fake” data generated in this way does not match the statistical properties of the real data and can thus yield dangerous misinterpretations and wrong assumptions about the nature of the real data. This in turn can result in implementation errors of ML algorithms, which could either lead to program crashes or statistical biases when the ML algorithm is trained with real data.

A promising way forward is thus the generation of realistic synthetic data. Synthetic data here refers to artificially generated data that retains at least some of the statistical properties of the original dataset while containing less information about a real person’s identity. In the context of healthcare, synthetic data can be created to mimic patient demographics, clinical characteristics, and other relevant features without exposing individual identities. Synthetic data can be generated via rule-based systems [8] or via generative artificial intelligence (AI) techniques [9]. There exist a wide range of applications of synthetic data, such as data augmentation [10], [11], [12], predictive modeling [13], [14], and the support of data scientists in the development of algorithms and tools [15], [16].

Although synthetic data is often considered a method to protect patient privacy, concerns have been raised in the literature about its effectiveness. Studies have shown that synthetic data may not always fully preserve privacy, raising questions about whether generative AI methods can truly fulfill this promise [17], [18], [19]. Due to problems such as an AI model overfitting of real training data, synthetic data can be used as an attack vector (e.g. through membership inference [20]) and thus retain the risk of leaking sensitive personal data [21], [22]. There exist numerous approaches to address this issue. One prominent approach is Differential Privacy (DP) [23]. While DP offers strong probabilistic privacy guarantees, it also has a strong impact on data utility. Utility will fluctuate with different degrees of DP guarantees [24], [25] and will generally decrease as the loss in privacy decreases [26].

This trade-off between synthetic data utility and privacy is an important topic in ongoing research. Brunelli et al. [27] propose a library and framework for evaluating privacy risks while ensuring statistical integrity of the data. They do, however, not provide an empirical evaluation of models or datasets to demonstrate the practical trade-offs between data utility and privacy in real-world scenarios. Latrup et al. [28] adopt a similar approach with their framework (Syntheval). They benchmark their framework using a hepatitis C dataset comprising 29 variables. Sarmin et al. [29] re-evaluate the privacy utility trade-off for anonymization-based sanitization against synthetic data generated by BayNet and PrivBayes using the Texas Hospital Inpatient Discharge Data Public Use Data File [30] (TEXAS) dataset. They found synthetic data to outperform sanitization enforcing k-anonymity in terms of balancing privacy and utility. Razi et al. [31] evaluate the utility of synthetic data generated by anyway conditional tabular generative adversarial networks (ACT-GAN) [32] as well as anonymized data using the PIMA Indian diabetes dataset [33]. They found that synthetic data showed higher utility than the differentially private and anonymized data.

Building on these previous works, our study aims to more systematically investigate the trade-off between synthetic data privacy guarantees and and the realism of synthetic data, i.e. fidelity. Unlike prior studies, which often evaluate single datasets or focus on limited metrics, our work conducts a comprehensive evaluation across three diverse datasets, including one that features complex, multimodal, and longitudinal data, which has not yet been evaluated in literature. We demonstrate how different privacy settings and different DP implementations can impact the fidelity of synthetic data on three different datasets covering different areas of healthcare, namely hospital discharge, clinical registries and longitudinal cohort studies. Additionally, we assess five distinct generative models, encompassing both models that incorporate differential privacy (DP) and those that do not, allowing for a robust comparison across approaches. Among these models we evaluate VAMBN [34], a method specifically designed to handle multimodal longitudinal clinical study data. Importantly, we also benchmark these models against traditional anonymization techniques, thereby providing a holistic view of the trade-offs between synthetic data generation and anonymization. To practically evaluate synthetic data fidelity and privacy in a visual and easy to understand manner, we have implemented SYNDAT as an open source framework^1^ for the evaluation and visualization of synthetic data, which is also available as a dashboard web application^2^ [35].

## 2. Methods

We generated synthetic data based on three different patient-level datasets using different models and privacy settings. We evaluated the resulting datasets in terms of their efficacy to preserve patient privacy and their data fidelity in terms of how closely they resemble the original data. Finally, we explored the practical utility of synthetic data for solving defined downstream tasks.

### 2.1 Selected datasets

We investigated synthetic data generated based on three different datasets covering medical research, medical routine as well as health economy:

1. The Texas Hospital Inpatient Discharge Data Public Use Data File [30] (TEXAS) has been released annually since 2006 by the Texas Department of State Health Services. It contains 18 economic hospital discharge data variables of 50000 patients from all state licensed hospitals in Texas, USA.
2. The Center for Cancer Registry Data of the Robert Koch-Institut (RKI) in Germany provides a dataset of all yearly new instances of new cancer cases in Germany [36]. The dataset consists of 18 variables for over 40000 individual records of patients with Glioblastoma Multiforme (GBM), a severe brain tumor. The dataset has previously been used in a statistical analysis investigating the effect of different treatment options (surgery, radiotherapy, chemotherapy) on overall survival [37].
3. The Alzheimer’s Disease Neuroimaging Initiative (ADNI) dataset [1] has been deployed for medical research purposes. It consists of features of different categories (clinical data, genetic sequencing data, volumetric imaging data) based on longitudinal screening data related to Alzheimer’s Disease (AD). The data was recorded over a period of 48 months at checkup interwalls of 6, 12 and 24 months for a total of 690 patients at different disease stages with a total of 239 variables.

### 2.2 Synthetization method

We generated synthetic data for the described datasets using five different generative AI models:

1. BayesianNet
2. PrivBayes
3. PateGAN
4. VAMBN
5. VAMBN using model training with DP (VAMBN-DP)

BayesianNet is a Bayesian Network implementation based on the implementation of DataSynthesizer [38], [39], an open source framework for AI-based synthetic data generation. PrivBayes is a DP version of the Bayesian Network model, which is also adapted from the same framework. The PateGAN model is based on its original implementation of Jordon et al [40]. PateGAN enforces privacy guarantees by adapting the Private Aggregation of Teacher Ensembles (PATE) [41] framework to the Generative Adversarial Network (GAN) architecture to generate differential private data. Variational Autoencoder Modular Bayesian Networks [34] (VAMBN). VAMBN is a generative modeling approach specifically designed for modeling longitudinal clinical study data with regular visit intervals [42]. It combines Bayesian Networks [43] and Heterogeneous Incomplete Variational Autoencoders [44] (HI-VAEs) to deal with heterogeneous data with potentially missing values without high risks of overfitting [34]. All models except VAMBN were natively supported by the framework of Stadler et. al. [17], which was additionally used for the evaluation of shadow model attacks (c.f. section 3.1). Notably, all methods except VAMBN ignore time dependencies within the data, i.e. BayesianNet, PrivBayes and PateGAN were just trained with plain tabular data as input. A minimal amount of preprocessing was applied to each dataset to prepare the data to be used as input for each generative model. Numerical data was normalized and categorical data was either ordinally or one-hot-encoded depending on the respective data type. We provide a list of used hyperparameters for each method and each dataset in the supplementary material in **Table S5**. We also provide supplementary configuration files for result reproduction of the VAMBN model in a separate Zenodo repository, see supplementary material.

In addition, we compared these data synthesization methods in terms of privacy risks against anonymization enforcing k-anonymity on demographic data as implemented in the NHSSanitizer tool by Stadler et al [45]. This was based on the idea that data synthesization can be seen as an anonymization process. The NHSSanitizer enforces k-anonymity [46] based on the demographic attributes of the original data. Sanitization is implemented by the imputation of missing values, outlier reduction for numerical values and rare categories as well as k=10 anonymization of demographic variables by record/row deletion. A list of the chosen Quasi Identifiers can be found in **Table S3** in the supplementary material. Full details on each model parameterizations are shown in **Table S5** in the supplementary material.

### 2.3 Differential privacy

We employed the (ϵ, δ)-DP framework originally proposed by Dwork et al. [23]: Let *A* be a randomized algorithm (here: a data synthetization algorithm) and 0 < ε, 0 < δ < 1. *A*: *D* → ℜ is said to respect (ε, δ) differential privacy, if for any two datasets *D*_1_, *D*_2_ ⊂ *D* that differ only in one single patient and for any output *S* ⊂ ℜ of the randomized algorithm, we have

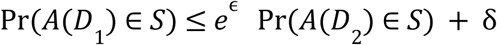

The parameter ε is oftentimes called the “privacy budget”, and δ constitutes a small relaxation of the guarantee enforced by pure ε-differential privacy, which corresponds to δ = 0. Abadi et al. [47] showed that it is possible to directly incorporate (ϵ, δ)-DP guarantees into the training of a neural network by clipping the norm of the gradient and adding a defined amount of noise to it, and this approach is practically followed in VAMBN-DP. The considered range of values for ε was chosen from the minimal possible ε that allowed at least one training epoch for VAMBN (high amount of noise added to gradient), up to 5 (low amount of noise added to gradient). This resulted in assessed ranges of ε = 1.27 to 5 (ADNI), ε = 0.45 to 5 (TEXAS) and ε = 1.02 to 5 (RKI) in step sizes of 0.1.

δ was fixed to 1/N, where N denotes the size of the dataset for each respective dataset, following a common recommendation by El Emam et al. [48]. While there is much discussion on the topic of choosing ε [49], configurations with values exceeding our tested ranges are common, specifically in situations where privacy aspects are not of utmost priority [50], [51].

### 2.4 Synthetic data fidelity

Synthetic data fidelity describes the degree to which synthetic data resembles the real data in terms of statistical properties. There is no widely accepted approach to measure the statistical similarity of two datasets, though. Intuitively, the statistical distributions of individual variables between synthetic and real data should match, and the correlation structure of variables should be preserved. Moreover, it may be expected that an ML classifier trained to differentiate between synthetic and real patient records should perform poorly.

Accordingly, we defined three main scores for each set of synthetic and real data, which are discussed in the following sections. For easier interpretation and better comparability, we normalized each score in the range of 0 to 100, where a higher score corresponds to a higher synthetic data fidelity. We included all scores in the SYNDAT tool for automatic, reproducible and equal assessments across datasets and synthetization methods.

#### 2.4.1 Marginal Statistical Distribution Similarity

One main goal of synthetic data generation is to produce data that should retain statistical properties of the original dataset. One of those properties is the statistical distribution of individual variables, i.e. the probability mass functions (PMFs) for discrete features and probability density functions (PDFs) for continuous features should resemble those in the real data. In practice, we estimated PDFs by binning values according to the Freedman rule [52]. We then calculated for each feature the Jensen-Shannon divergence (JSD) of real versus synthetic value distributions and computed the average over all features. The JSD is bounded by 1; a low value can be interpreted as having a similar statistical distribution, with a value of zero meaning that two distributions are equal. We normalized this divergence measure into the range of [0,100] as a score to make it easier interpretable:

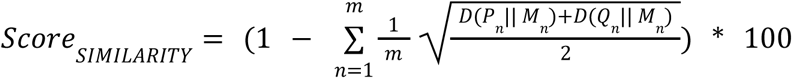

*D* is in this formula the Kullback Leibner (KL) divergence and *M* is the pointwise mean of the distributions *P* and *Q* of the real and synthetic feature *n*, respectively.

#### 2.4.2 Correlation Structure

Additionally to having similar marginal distributions of individual variables, synthetic data should also show a correlation structure similar to real data. Importantly, having similar marginal distributions does not imply that statistical dependencies between variables are kept. To quantify this, we specifically used the Spearman rank correlation to capture potentially nonlinear monotone statistical dependencies between variables in both the real and synthetic datasets. We thus compared the Frobenius norm of the difference of the correlation matrices of the real and synthetic datasets, respectively, to the Frobenius norm of the correlation matrix of the real data. After thresholding at zero, this quantity may be interpreted as the relative error of the correlation structure of the synthetic data. We subsequently rescaled the relative error to a score between 0 (low similarity) and 100 (high similarity):

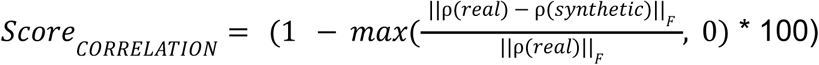

While in theory, the correlation quotient can exceed 1, this typically occurs when there is either minimal or no correlation in the real data compared to a higher correlation in the synthetic data, or when mostly opposite correlations structures are present in real and synthetic datasets. As both of these scenarios show distortions in the synthetic data that imply poor data quality in terms of representation of the real data, we consider correlation coefficients exceeding 1 with the lowest possible score of zero.

#### 2.4.3 Discrimination Ability

A common way to assess the similarity of real and synthetic data is to train a machine learning classifier to differentiate between real and synthetic records. If the synthetic data closely resembles the original data in terms of marginal statistical distributions and correlation structure, a trained machine learning classifier should only achieve a low prediction performance when trying to differentiate between the real and synthetic data points. We here trained a *Random Forest* (RF) classifier to differentiate between the two classes of real and synthetic data points within a 5-fold stratified cross-validation scheme, ensuring that each training fold maintained the same proportion of real and synthetic subjects. In case of the longitudinal ADNI data, we concatenated features from all visits but omitted all with more than 10% missing values to avoid a trivial differentiation between real and synthetic cases by the rate of missingness. We calculated the average *Receiver Operating Characteristic* (ROC) *Area Under Curve* (AUC) for all five test splits to evaluate the classifier performance:

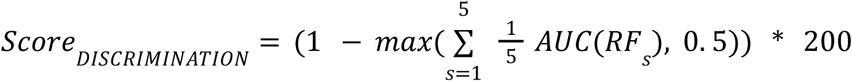

We scale the inverse of the AUC scores by the factor 200 since the optimal AUC, which corresponds to a prediction performance at chance level, is in this case 0.5. In case of an AUC under 0.5 we also report an optimal score of 100.

We deliberately evaluate three different evaluation scores to assess the fidelity of synthetic data that reflect different statistical aspects. While, for instance, the average JSD and even the relative error of the correlation matrix may be low, a single feature that is not well modeled may result in a high prediction performance of the RF classifier, as the classifier learns to discriminate based on that single feature. In this case the *Discrimination Score* would strongly underestimate the synthetic data fidelity, while the *Similarity* and *Correlation Scores* would slightly overestimate since they are effectively based on an average of over all features. It is therefore important to inspect different facets of the synthetic data to adequately assess its overall fidelity.

### 2.5 Synthetic data utility

In addition to fidelity, practical utility of synthetic data for specific data analysis tasks is a further important aspect. In this paper we restrict ourselves to synthetic data generated by VAMBN for this purpose, because VAMBN is applicable to static as well as longitudinal data and showed superior fidelity of generated data (see Results section). Due to SanitizerNHS also archiving high fidelity results (c.f. section 3.1), we added it as an additional baseline next to the real data. To explore the utility of synthetic data generated by VAMBN, we considered the following tasks associated to each of the investigated datasets:

- Task 1 (TEXAS): Prediction of illness severity and mortality risk. We train a RF classifier to differentiate between minor/moderate (groups 1-2) and major/extreme (groups 3-4) illness severity or mortality risk.
- Task 2 (RKI): Prediction of overall survival
- Task 3 (ADNI): Prediction of cognitive impairment (cognitively normal (MMSE > 23) vs cognitively impaired (MMSE < 24)) at study baseline

For tasks 1 and 3 a Random Forest classifier was trained, while for task 2 a Cox regression model was employed. In all cases models were trained on synthetic data and tested on real data. The same number of synthetic as real data instances were produced, and the synthetic data generation and training process was repeated 10 times to account for the randomness in the synthetic data generation. All models were evaluated on a held-out test set consisting of 20% of the real data. Details on the variable selections are shown in the supplementary material. Performance metrics for tasks 1 and 3 models were AUC-ROC, whereas models in task 2 were evaluated via Harrel’s C-index [53].

### 2.6 Privacy assessment

#### 2.6.1 Membership Inference

In recent years, it has been repeatedly shown that synthetic data is susceptible to membership inference attacks, i.e. attacks that try to reveal whether a real patient’s record is included in a synthetic dataset [45], [54]. The risk of linkability addresses the ability of an attacker to link published, protected data records to a single record, or group of records, thus leading to inference of sensitive information or re-identification of the target [55].

One approach to assess membership inference risk using a privacy game between an attacker and a data publisher [45], [56]. Stadler et. al. proposed a framework^3^ based on the idea from Shokri et al. [54], that treats the synthetic data generator as a black box and performs iterations of membership inference attacks on selected target records. During such a “game”, the attacker selects multiple sample datasets from a published, non-protected reference dataset with a similar statistical distribution as the original, non-published dataset. These samples explicitly include and exclude a target record. Then, a set of so-called shadow models is trained using the synthetization approach as black-box. More specifically, we here trained a Random Forest on the synthetic data to predict whether a given target record was included in the original real training dataset. The risk of membership inference is then expressed as privacy gain, holding the assumption that membership inference is always successful, if a record of the original dataset is provided to the classifier:

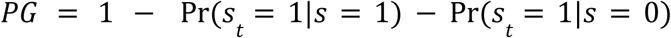

Here Pr(*s_t_* = 1|*s* = 1) is the probability of a membership prediction under the condition of an actual appearance of the target record in the training data. Hence, the privacy gain is expressed as a relationship between the classifiers true positive and false positive rate. This allows to show an actual privacy leak as the difference between these values, i.e. under ideal conditions *PG* should be 1. The accuracy of these classifiers is of lower importance.

For the selection of suitable target records, we utilized an outlier score that calculates the normalized Euclidean distance between records and the dataset centroid [57]. For this purpose, each categorical variable was encoded by its PMF. We then selected the 3 strongest outliers as well as 3 records near the centroid for our assessment of residual risks.

#### 2.6.2 Singling-Out and Inference Risks

In addition to shadow model attacks inferring membership, we considered the privacy risk evaluation framework Anonymeter [58] introduced by Giomi et al. [18]. The authors proposed to calculate privacy risks as the percentage of successfully guessed information on records of the real data using the synthetic data. We evaluate two risks:

1. **Singling-Out Risk**: An individual can uniquely be identified from a dataset, without using direct identifiers.
2. **Inference Risk:** Information about an individual can be deduced or inferred from the data, even if the data does not explicitly include that information.

For our experiments, we evaluated these risks using a 80/20 train/test split of the generated data. The test split was used as a control set, which Anonymeter uses to evaluate the excess privacy risk. In addition to the main attack executed on the real data and control attack executed on the control data, Anonymeter will perform a baseline attack as a sanity check that models an attacker that tries to infer information by random guessing. The authors implement this as a mechanism to avoid risk underestimation in case of a weak attacker. We additionally highlighted computed results where the sanity check of Anonymeter failed.

For the Singling-out Risk, we chose to evaluate multivariate attacks only, as univariate attacks generally performed poorly in comparison to the baseline model. We additionally repeated attacks which reported that performed worse or equally bad as the baseline model while averaging results for valid runs to make our results more robust.

## 3. Results

### 3.1 Fidelity of the synthetic data

We evaluated data fidelity of each dataset for every model based on the three fidelity scores that we introduce in section 2.4. A summary of all computed scores can be found in **Table 1**. We denote the *Discrimination score* as **M1**, the *Distribution score* as **M2** and the *Correlation score* as **M3**.

**Table 1:**
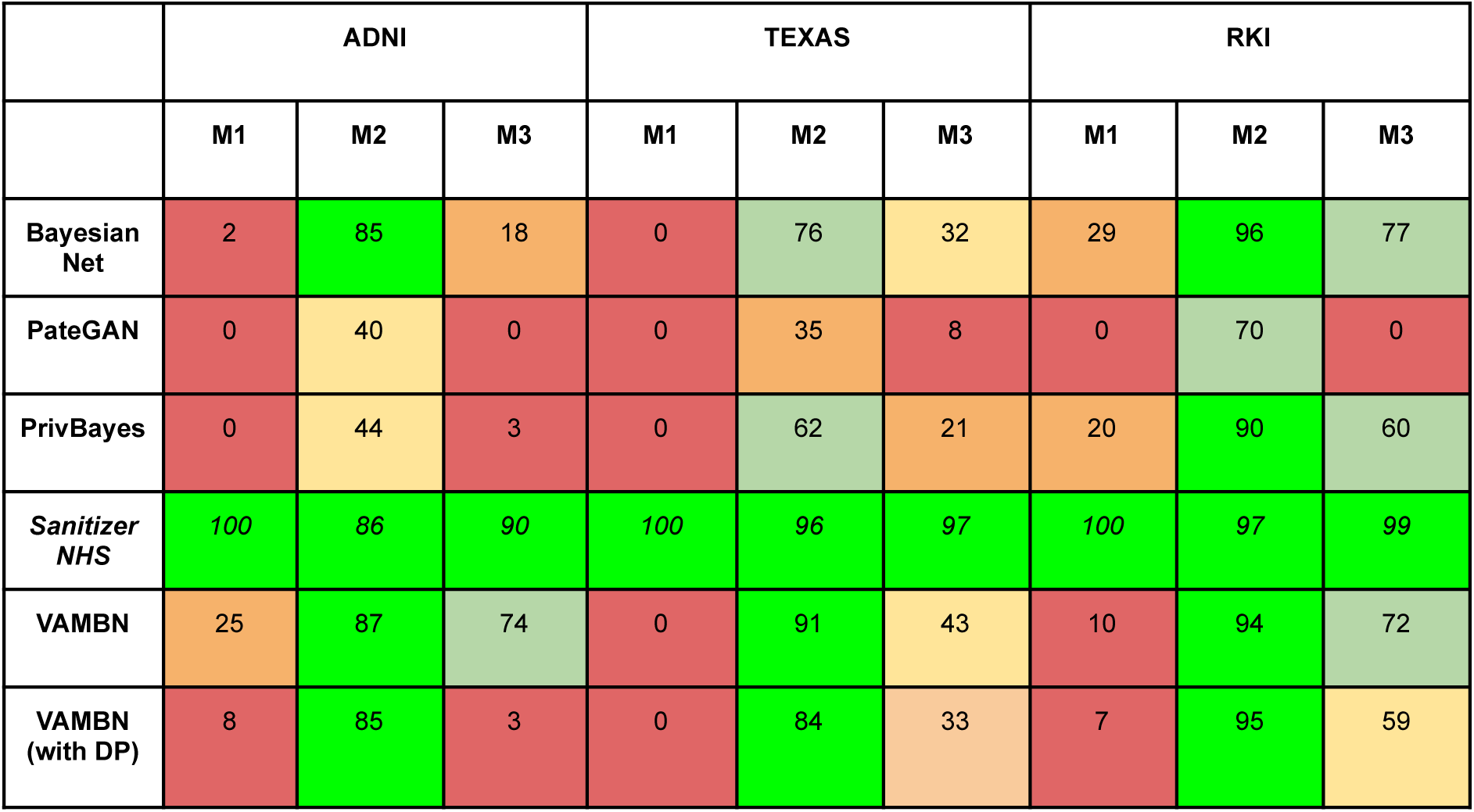
Three calculated fidelity metrics (M1: Discrimination, M2: Distribution, M3: Correlation) for all techniques and datasets. SanitizerNHSk10 is an anonymization method of demographic features, but not a synthetic data generation approach. It is only shown for comparison reasons.

For the ADNI dataset, SanitizerNHS demonstrates consistently high fidelity across all three metrics (M1: 100, M2: 86, M3: 90), outperforming all synthetic generation models. Among the synthetic data generation methods, VAMBN achieves the highest overall performance (M1: 25, M2: 87, M3: 74) especially in terms of Distribution and Correlation Score. PateGAN and PrivBayes exhibit notably poor results in Discrimination and Correlation, with PateGAN yielding zeros for both M1 and M3. The inclusion of differential privacy in VAMBN (with DP) notably removes the model’s effectiveness in preserving correlation (M3: 3) but has a marginal impact on the Distribution Score.

The performance patterns observed in the TEXAS dataset are generally consistent with those in ADNI. SanitizerNHS generally preserves the original data well, with high scores in all three metrics. Distributions are well preserved for both DP and non-DP VAMBN, while the preservation of correlations for all synthetic generation models are generally worse compared to ADNI. Discrimination Scores (M1) are zero for all synthetic generation methods, which can be attributed to the fact that correlation preservation was generally low for all models. PateGAN performs poorly for all computed metrics. All models implementing DP again lose performance in terms of correlation preservation, while not being as pronounced as in the ADNI dataset.

In the RKI dataset, SanitizerNHSk again maintains high fidelity. The BayesianNet model performs well, particularly in Distribution (M2: 96) and Correlation (M3: 77), while VAMBN maintains solid performance in Distribution (M2: 94) and Correlation (M3: 72). The addition of differential privacy in VAMBN (with DP) leads to a similar trend of reduced performance in Correlation (M3: 59), though its Distribution (M2: 95) score remains high. PrivBayes shows moderate fidelity with a strong Distribution score (M2: 90), while its Correlation Score is similarly reduced as in the VAMBN DP model.

Overall, in terms of DP, two trends can be observed: Across all datasets, the introduction of DP in the VAMBN model leads to a significant reduction in correlation preservation (M3). This trend is most evident in the ADNI dataset, where the Correlation Score drops from 73 in the standard VAMBN to 3 in VAMBN with DP. Similar reductions are observed in the TEXAS and RKI datasets, where M3 falls from 52 to 36 and from 71 to 58, respectively. The same trend can be observed for the Bayesian Network based models, where we see an average decrease of the Correlation Score of 21 points when comparing PrivBayes with BayesianNet.

These findings indicate that the noise introduced by DP mechanisms strongly affects the model’s ability to maintain relationships between variables, which is critical for some analytical tasks that rely on these correlations.

In contrast, the Distribution Score (M2) is relatively robust to the inclusion of DP. In the ADNI, TEXAS, and RKI datasets, VAMBN with DP maintains high M2 scores (85, 84, and 94, respectively), only slightly lower than the non-DP version of the model. This suggests that while DP significantly impacts the model’s ability to preserve variable correlations, it has a minimal effect on preserving the overall distribution of the data.

While results for correlations and feature distributions appear generally consistent across the datasets, performance of the synthetic data generation models in terms of discrimination (M1) shows notable variation. Other than in cases of generally low preservation of correlations as seen in the TEXAS dataset, low Discrimination Scores also occur for single outlier features due to modeling artifacts which are then learned by the classification model.

We illustrate this example for the case of non-DP VAMBN for the ADNI dataset; while both the M2 and M3 is high due to features are well modeled *on average,* a SHapley Additive exPlanations (SHAP) [59] analysis (see Supplementary **Figure S2**) on a model trained to discriminate real and synthetic data showed a single feature with significantly high impact on the model prediction. Further investigation of the feature (MMSE) showed synthetic values outside of the real data range (**Figure S1**), which is by clinical definition limited at a maximum of 30. These constraints cannot be derived from observed data alone and would require additional post-processing steps or the design of customized loss functions during model training based on available background knowledge.

### 3.2 Utility of synthetic data

Anonymized data using SanitizerNHS showed high utility for all three experiments, which was expected due to its high data fidelity (see section 3.1). For the ADNI data set, the anonymized data showed a lower utility than the synthetic data generated by VAMBN (AUC 0.80 vs 0.85), likely due to the high amount of deleted records, impacting the training of the classifier.

For ADNI and RKI datasets, all models trained on synthetic data generated by VAMBN also demonstrated a prediction performance close to that achieved on real data (ADNI: AUC 0.88 on real vs. 0.85 on synthetic data; RKI: C-index 0.73 on real vs. 0.72 on synthetic data). However, for both models trained on synthetic TEXAS data a substantial decline in performance was observed. The results of all four prediction tasks are shown in **Figure 1**.

**Figure 1:**
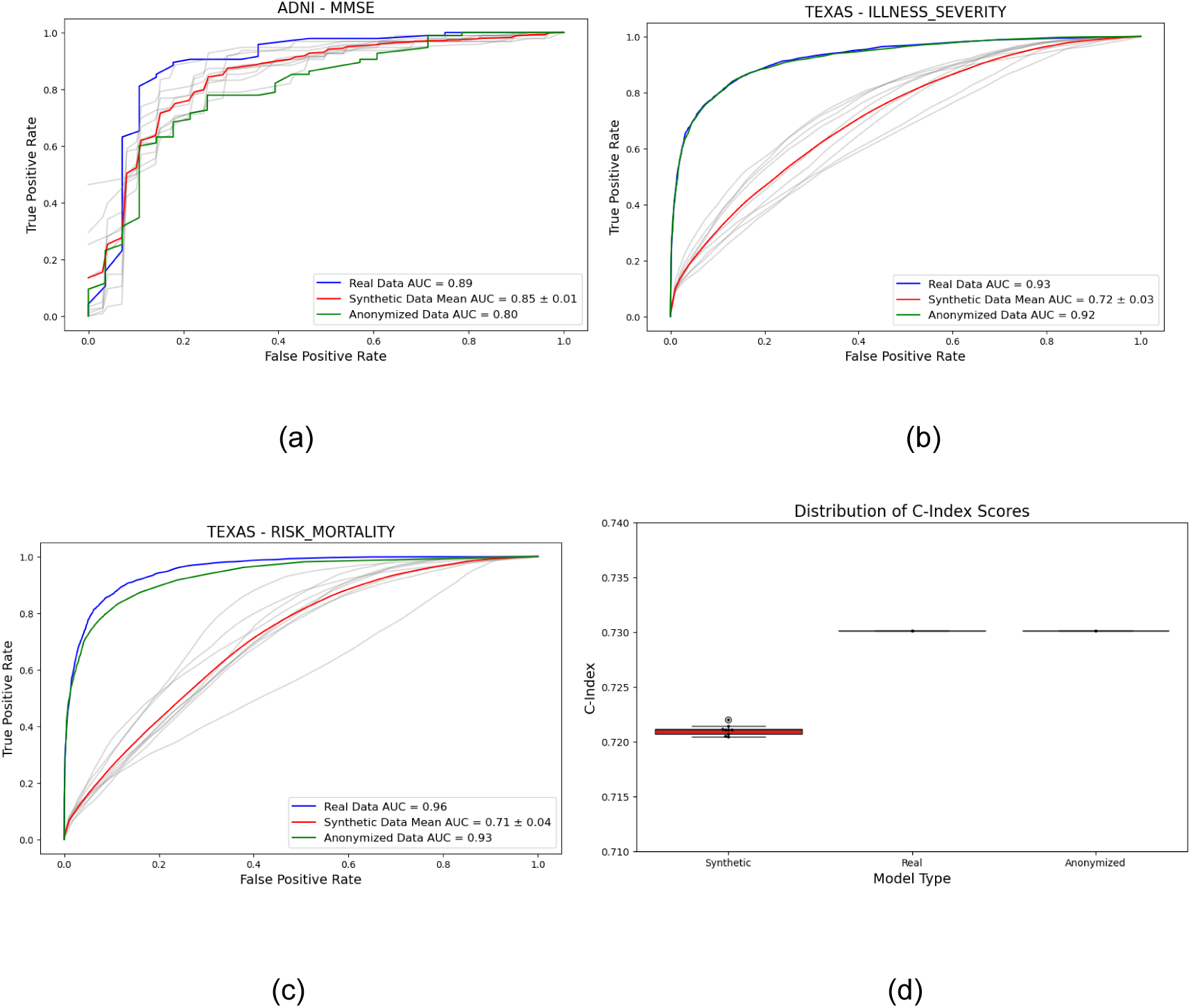
Utility evaluation results for each dataset. (a) Prediction of (MMSE) using ADNI. (b) Prediction of illness severity using TEXAS. (c) Prediction of mortality using TEXAS. (d) Prediction of survival probability using RKI.

To explain the substantial decline in model performance after training on the synthetic TEXAS data, we computed SHAP values for the model trained to predict mortality risk (**Figure 1c**, **Figure 2**). Based on this analysis, the features “illness severity” and “patient age” were found to have the biggest impact on model predictions. While we found that marginal distributions of synthetic TEXAS data for these two features align well with the real data (**Figure 3**), the pairwise correlations differ substantially (**Figure 4**). This agrees with the results of our computed fidelity metrics (**Table 1**). While VAMBN performed well in terms of its distribution score (**Table 1, M2**) for all three evaluated datasets, we saw a lower Correlation Score of 52 for TEXAS compared to a Correlation Score of >70 for both ADNI and RKI. This suggests that out of the three evaluated scores, a high Correlation Score is likely to be a good indicator for data fidelity. While the Discrimination Score appears useful to identify single features that are not aligned with the original data, their effect seems negligible for the overall data utility. We can see that even in cases of a low Discrimination Score (i.e. VAMBN on RKI), a high utility can be obtained. In fact, we can observe on the example of ADNI and its deviating single feature (see **Figure S3**), that data utility may be mostly unaffected (as evident in the utility results **Figure 1 (a)**), if the correlation structure (high MMSE, i.e. low incidence of AD) is still maintained in the synthetic data, even though the actual value ranges do not entirely match the ranges of the real data.

**Figure 2:**
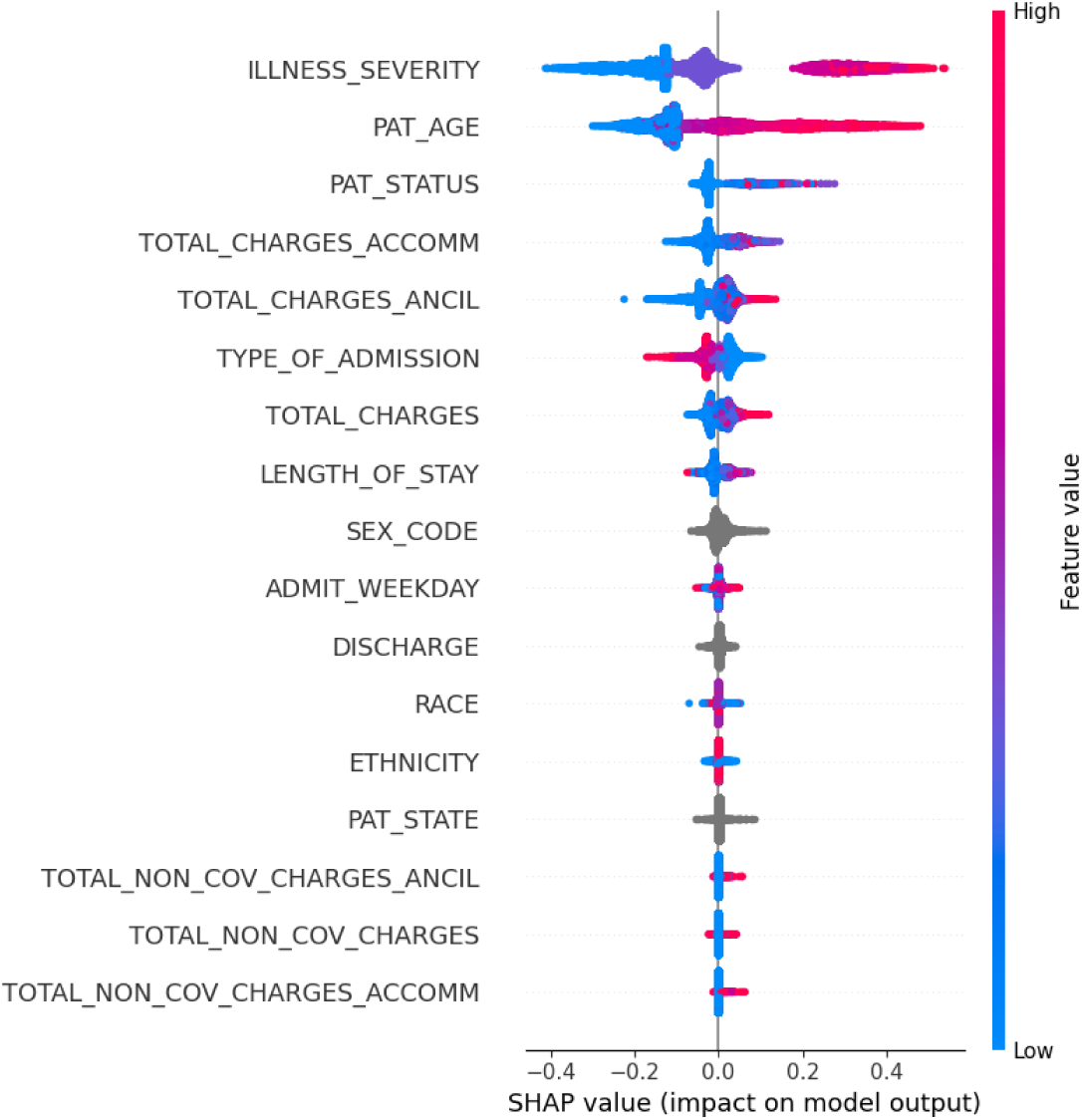
SHAP values for a model trained on real data to predict mortality risk. The two features with the biggest impact on mortality risk were found to be high illness severity as well as a high patient age.

**Figure 3:**
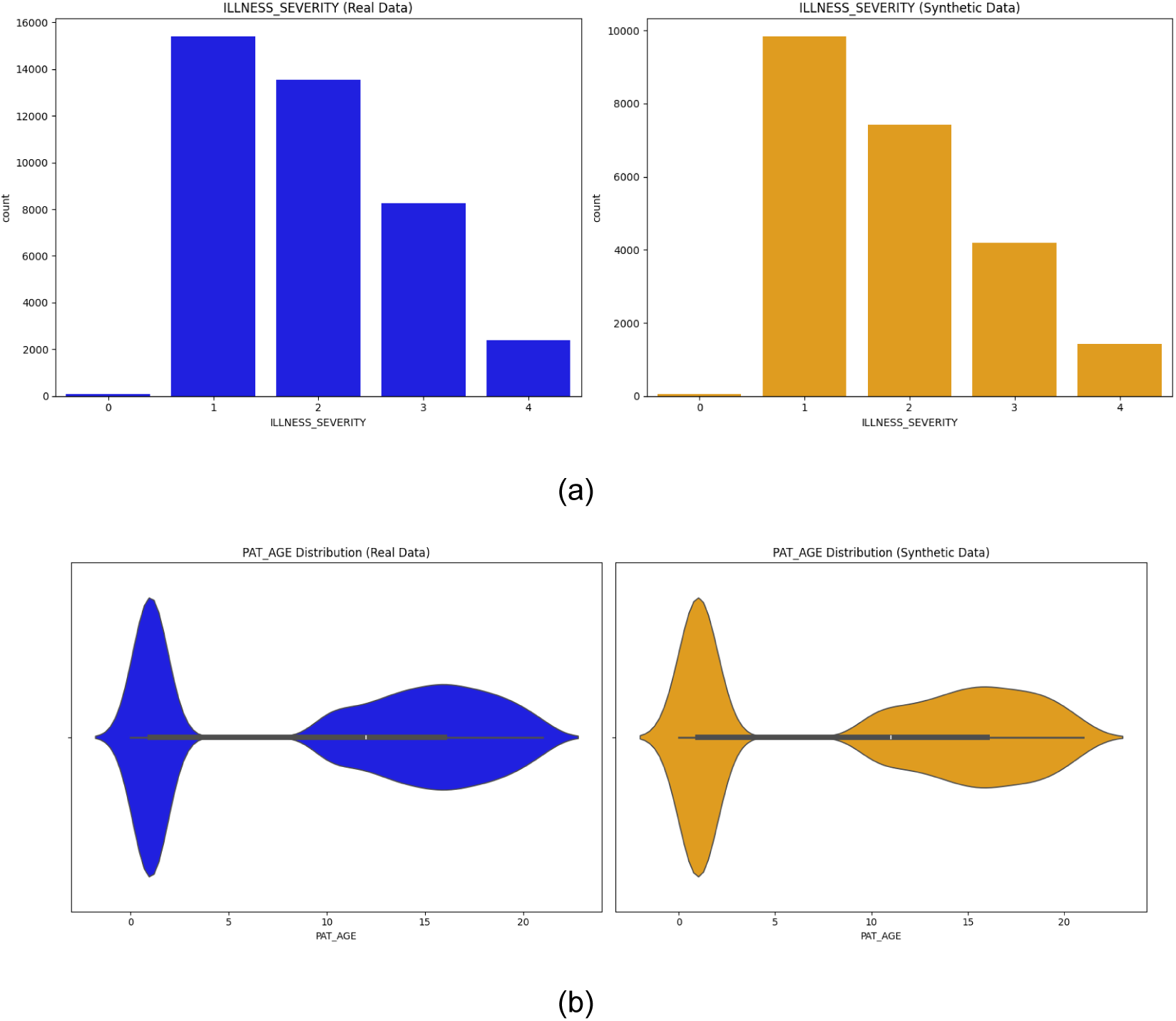
Real and synthetic marginal distributions for the features of illness severity (a) and patient age (b) for real and synthetic TEXAS data.

**Figure 4:**
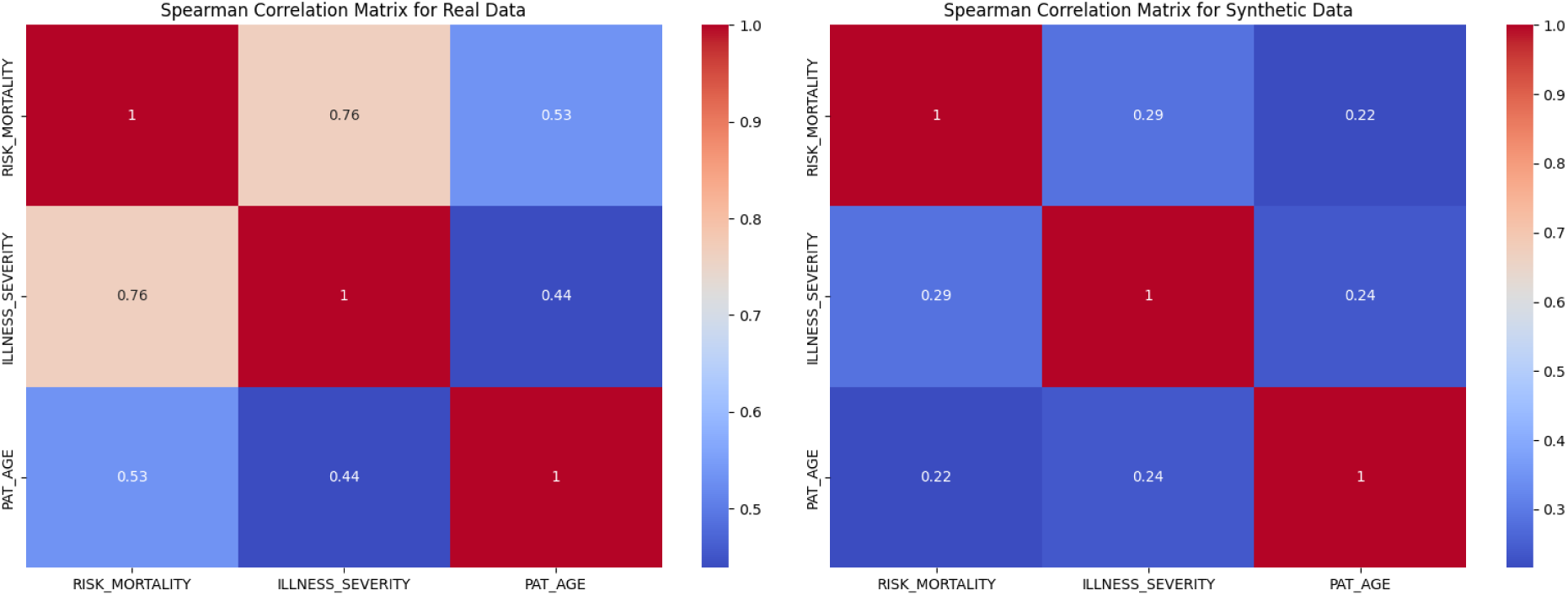
Pairwise feature correlations between illness severity, patient age and risk mortality for real and synthetic TEXAS data.

### 3.2 Privacy of the synthetic data

#### 3.2.1 Membership Inference

Privacy evaluation of the ADNI dataset was slightly limited since PateGAN as well as the NHSSanitizer were unable to generate a sufficient number of records due to the subsampling of the relatively limited real data (n=580), which is required for shadow-model attacks. Apart from this aspect the PG score averaged around 1 and thus indicated no obvious membership inference risks (**Figure 5 (c)**). However, a high variance across different runs of the attack game was observed, indicating potential higher risks for individual targets. No statistically significant differences across the tested synthetic data generation methods could be found.

**Figure 5:**
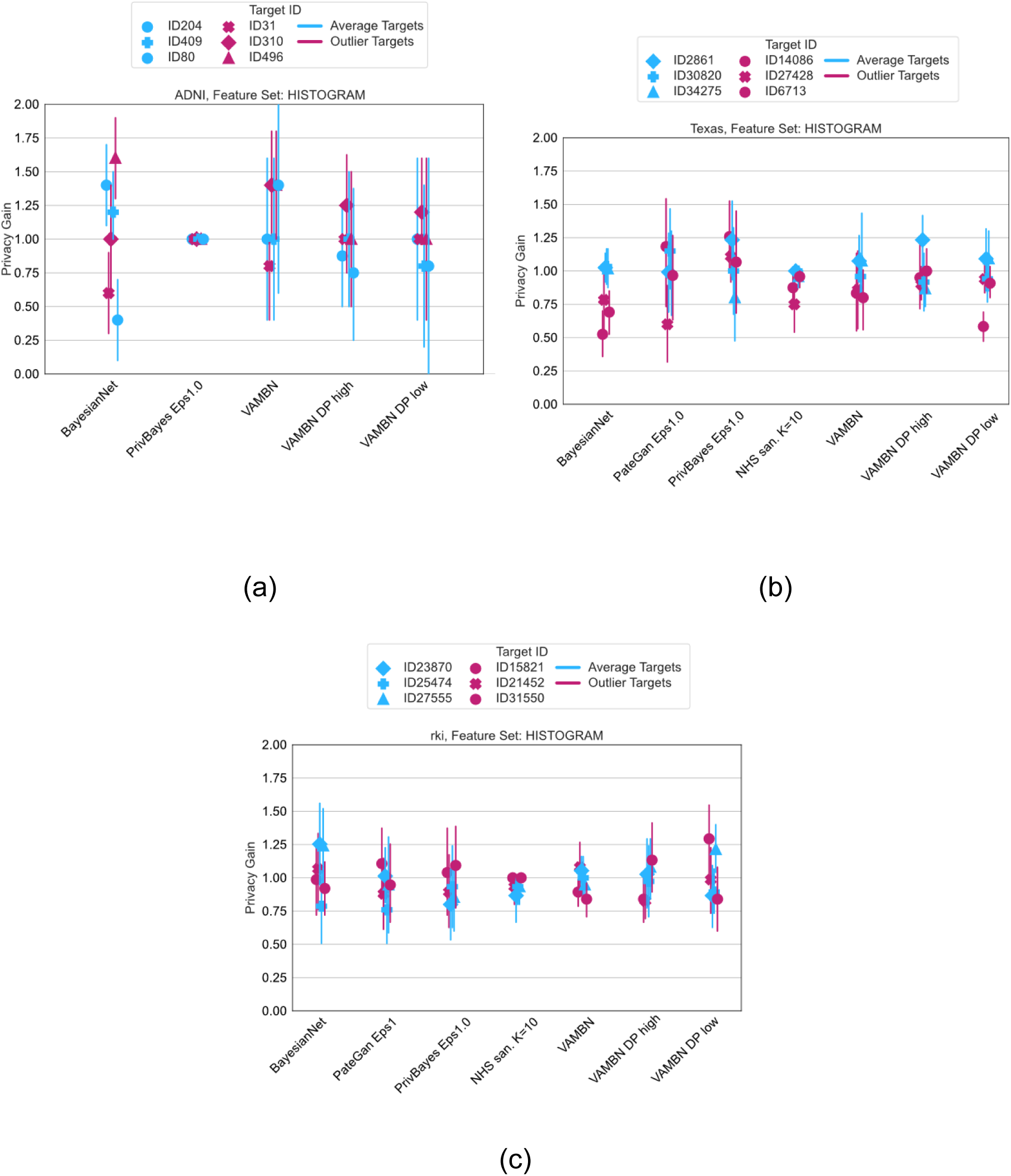
Risk of membership inference expressed by PrivacyGain for the ADNI (a), TEXAS (b) and RKI (c) dataset using shadow model attacks. A PrivacyGain of 1 indicates an equal amount of correct guesses on target and non-target included data samples, hence perfect protection.

For the TEXAS dataset, the PG score averaged around 1 for typical records, with more significant deviations observed in outlier targets, particularly in synthetic data generated by BayesianNet, VAMBN and VAMBN with low DP. Compared to the VAMBN implementation without DP, the higher protection levels in the dataset may thus improve defenses against membership inference and reduce information leakage.

Similarly, the RKI dataset showed a consistent PG score around 1, indicating good average protection. Notably, there was no significant difference between outlier and average target records, suggesting that the original, unprotected dataset may already offer robust protection against membership inference. For detailed visualizations, see **Figure 5**.

#### 3.2.2 Singling Out and Attribute Inference Risks

The Anonymeter framework provides standardized risk scores between 0.0 and 1.0, where higher values indicate higher residual privacy risks. We used 10 iterations of Anonymeter to provide stable results (**Table 2**). Notebly, Anonymeter failed the sanity checks (c.f. section 2.6.2) for singling out attacks for all models on the RKI and TEXAS datasets as well as for the PateGAN model on the ADNI dataset. The reason might be that attack success rates were lower or equal than pure chance (c.f. section 2.5.2).

**Table 2:** Calculated Risk for Singling Out and 95% confidence interval (brackets) for all 3 datasets and all evaluated models. In cases annotated with (*), Anonymeter attacks were deemed not successful because neither baseline nor actual attack resulted in any successes.

| Singling Out Risk | ADNI | TEXAS | RKI |
| --- | --- | --- | --- |
| **BayesianNet** | 0.11 (0.02) | 0.00 (0.00, 0.00)* | 0.00 (0.00, 0.00)* |
| **PateGAN** | 0.00 (0.00, 0.00)* | 0.00 (0.00, 0.00)* | 0.00 (0.00, 0.00)* |
| **PrivBayes** | 0.06 (0.01) | 0.00 (0.00, 0.00)* | 0.00 (0.00, 0.00)* |
| **SanetizerNHS** | **0.25 (0.03)** | 0.00 (0.00, 0.00)* | 0.00 (0.00, 0.00)* |
| **VAMBN** | 0.11 (0.02) | 0.00 (0.00, 0.00)* | 0.00 (0.00, 0.00)* |
| **VAMBN-DP high** | 0.12 (0.02) | 0.00 (0.00, 0.00)* | 0.00 (0.00, 0.00)* |
| **VAMBN-DP low ($\epsilon=5$)** | 0.11 (0.02) | 0.00 (0.00, 0.00)* | 0.00 (0.00, 0.00)* |

Overall, residual privacy risks for singling-out based on multivariate analysis showed no identifiable risks for the TEXAS and RKI datasets across all tested methods and only few successful singling out attacks for the ADNI dataset. For the latter case, we observed comparable results for the BayesianNet, PrivBayes as well as all VAMBN variants (all risk scores <= 0.12). No specific advantage was observed for DP techniques. The highest residual risk scores of 0.25 was found for the NHSSanitizer, which only performs anonymization of demographic features.

Regarding attribute inference risks estimated by the Anonymizer framework, we found residual risks scores below 0.2 for all synthesization techniques for all datasets (**Figure 6**). Again, highest risks were found for the NHSSanitizer on TEXAS with values of up to 0.8 indicating a high likelihood that the anonymized data could be used to infer values of some attributes of real patients.

**Figure 6:**
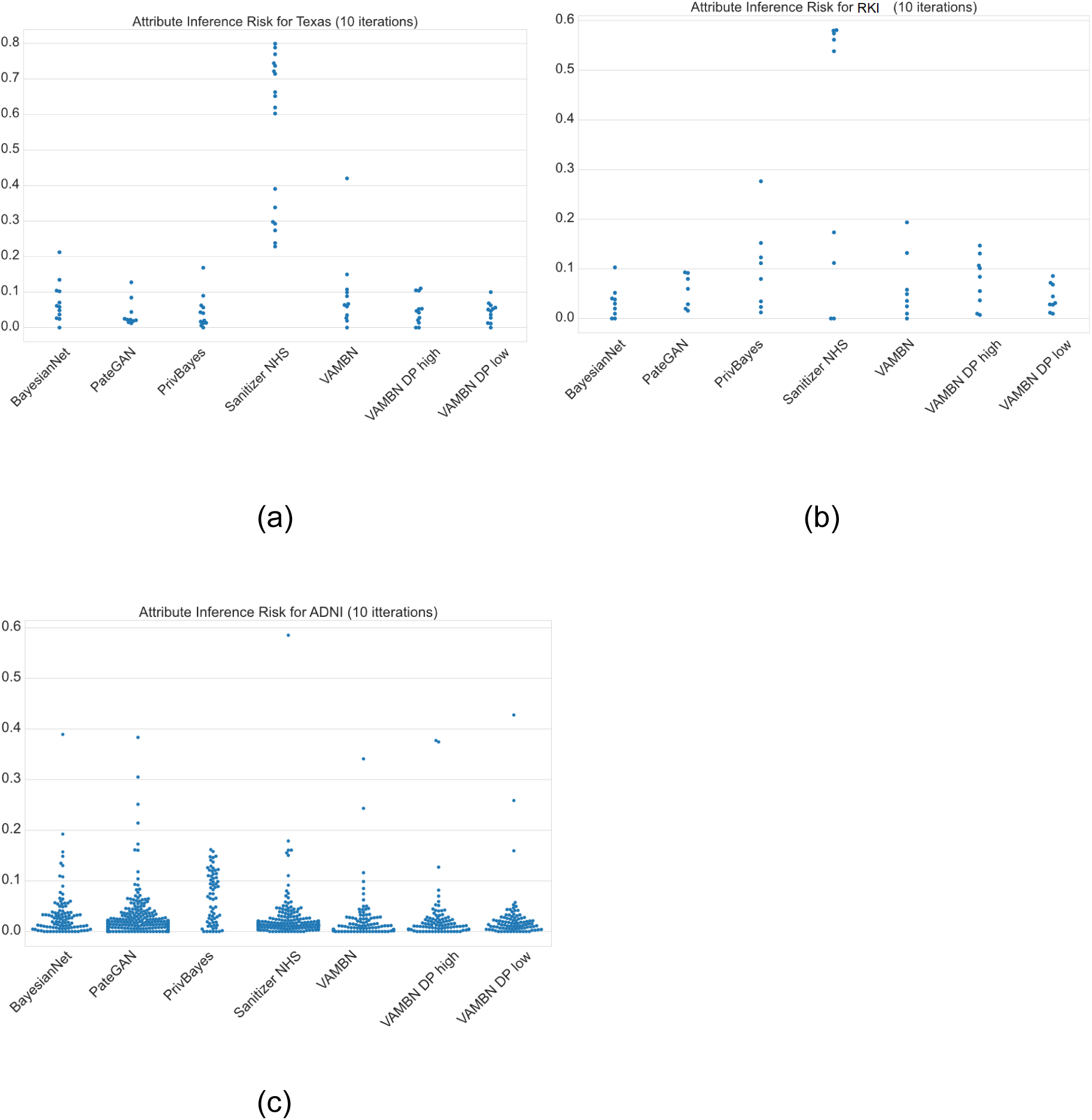
Attribute Inference Risk over all features averaged over 10 iterations for TEXAS (a), RKI (b) and ADNI (c)

## 4. Discussion

This study aimed to evaluate synthetic data fidelity of each dataset based on the proposed metrics for both DP and non-DP models and assess the potential trade-off in terms of privacy preservation and data utility. To perform a broad range evaluation of data fidelity, we chose to assess three different fidelity scores, focussing on different aspects of the synthetic data.

Modern generative AI methods, particularly VAMBN, demonstrated the ability to generate patient-level synthetic data with high fidelity. Synthetic data generated by this method can be leveraged to train machine learning models that achieve predictive performance comparable to those trained on real-world data. However, as shown by the results from the TEXAS dataset, even minor discrepancies between synthetic and real data can lead to significant variations in model performance. Therefore, it is crucial to conduct thorough quality assessments of synthetic data before utilizing it for downstream analyses or applications.

Based on the results of our utility evaluation, we identified the Correlation Score to be the most important factor to preserve utility in synthetic data out of the tested scores. While generative models such as VAMBN were generally able to preserve feature correlations in a complex and multimodal dataset such as ADNI, the addition of differential privacy to the model was detrimental to correlation structures and resulted in a near complete loss of feature correlation, thus making the synthetic data effectively useless for any down-stream tasks. While we also found a significant decrease in Correlation Scores in the structurally simpler RKI and TEXAS datasets, the negative effect was less pronounced, thus potentially preserving a higher degree of utility in DP settings for less complicated underlying modeled data.

Although our study found that differential privacy (DP) significantly affects feature correlations for all tested implementations, its impact on the marginal distributions of individual features is minimal. This indicates that, although certain implementations of DP may not be optimal for model training, they can still provide valuable insights at a univariate feature level and yield general summary statistics of the original dataset.

While we found that our proposed scores - and specifically the *Correlation Score* - translate well into actual synthetic data utility, they also have their limitations. While the proposed scores to evaluate synthetic data fidelity provide a good first impression, they are still dependent on the structure of the real data. For instance, the *Feature Correlation Score* will be less informative for data in which features show little to no correlation. Variability in data types, such as categorical versus continuous variables, can further impact the results - especially the *Distribution Similarity* score - and depend on the chosen encoding of categorical features. Lastly, we found that the computed *Discrimination Score* may in most cases be overly pessimistic, as a Random Forest classifier was able to predict for all cases rather reliably whether a data point was a real or synthetically generated one, even if both the statistical distributions and correlations were relatively well reproduced. Future research should thus focus on extensions or modifications of the proposed scores to make them more robust and more generally applicable across different data structures, including, e.g., unstructured imaging or text data. Furthermore, we would like to extend our analysis in the future to include more recent algorithms for DP synthetic data generation, such as AIM [60], to explore their capabilities in enhancing feature correlation preservation in comparison with the already evaluated methods and to assess their overall impact on data utility and privacy.

In line with our fidelity and utility assessments, we further evaluated the privacy-preserving capabilities of each model using shadow model attacks, alongside assessments of Singling Out and Inference Risks through the Anonymeter framework. Neither approach revealed any significant privacy concerns across the tested data synthesis techniques. Notably, we observed no substantial differences in privacy risks between synthetic data generation methods with and without DP, raising questions about the practical benefits of incorporating DP in AI model training. At the same time a key limitation we identified is the inability of shadow model attacks to effectively handle high-dimensional datasets with relatively small sample sizes, as exemplified by the ADNI dataset (**Figure 5c**). Additionally, the high computational costs associated with shadow model attacks further challenge their practicality in real-world applications.

Our privacy analysis via Anonymeter of k-anonymization using SanitizerNHS revealed high degrees of privacy leakage in terms of Inference risks for RKI and TEXAS as well as Singling Out Risks for ADNI. While the method was able to preserve fidelity to a high degree, the risk analysis using Anonymeter highlighted the susceptibility of SanitizerNHS against singling out and attribute inference attacks, indicating that it does not provide a good risk/utility trade-off. Both our experiments and former research [61] have shown that DP provides better privacy protection concerning these risks.

Our findings generally support the results we have seen in previous studies [29], [31]. Synthetic data is favorable in terms of the utility versus privacy trade-off when compared to anonymized data. DP synthetic data exhibits reduced utility when compared to regular synthetic data - and will not necessarily provide significantly better privacy protection as regular synthetic data can already provide sufficient privacy guarantees.

Overall, our findings emphasize the delicate balance between data fidelity, utility, and privacy in synthetic data generation in all evaluated methods, highlighting the importance of tailoring privacy-preserving methods like VAMBN with differential privacy to the specific characteristics of the dataset, while ensuring that fidelity metrics are carefully chosen to reflect both the structure and intended use of the data.

### Conclusion

Synthetic data bears strong potential for facilitating data sharing in healthcare. If sufficiently realistic, synthetic patient-level data may be used for (pre-)training AI/ML models [62], for augmenting real data, including oversampling of minority classes, and for generating synthetic control arms for clinical studies, which could be shared between organizations. At the same time, the intricate trade-off between fidelity and privacy of synthetic patient data is a critical area of study and recent work has highlighted that this is done too seldomly [63]. Our investigation underlines the complexity and nuances involved in assessing this balance, and presents a practical approach for doing so.

Our results show that when data fidelity is maintained, synthetic data can be successfully used for training predictive modeling with a similar model performance as if one were to train on real data. However, we also found that when data fidelity is compromised - especially in terms of preservation of correlation structure - it has a detrimental effect on data utility. This was specifically the case when applying DP respecting model training. Our findings thus suggest that implementations of DP are not yet practically useful for generating realistic synthetic patient data in complex datasets and highlight the need for further research to enhance the practicality and effectiveness of DP preserving techniques in this domain.

Both tested privacy assessment frameworks revealed no obvious privacy breaches due to synthetic data generation by any of the tested techniques, even without DP. Of course, this does not imply that according risks do not exist, but just that neither Anonymeter nor shadow model attacks were able to identify them. In this regard, our analysis highlighted obvious shortcomings of shadow model attacks, specifically in case of high-dimensional longitudinal clinical study data. Future research should just explore better approaches to assess the privacy of patient-level synthetic medical data, specifically considering the longitudinal aspect of many of those datasets.

Altogether, the absence of standardized guidelines and best practices for data sharing, data protection and synthetic data privacy evaluation in healthcare can lead to significant inconsistencies. These inconsistencies might result in varying conclusions by different stakeholders about the fidelity, utility and security of synthetic data. Without a unified approach, different healthcare institutions may thus implement different strategies, making it difficult to ensure that patient data is treated according to the same standards. Hence, there is a need for clear technical guidance with regard to synthetic patient data fidelity, protection and privacy assessment.

## Limitations of the study

The study evaluates three patient-level datasets, which may not fully represent the diversity of medical patient-level data. The findings might therefore not generalize to all types of patient level data.

The study assesses five different models for data synthetization and anonymization with and without the use of DP. With the large number of models available and the frequent development of new approaches, the study does not cover the whole spectrum of existing models. Other models could show different privacy-utility trade-offs.

## Data Availability

This study did not generate new unique reagents.
The data supporting the findings of this study are available from the respective data holding organizations (c.f. section 2.1). Researchers interested in accessing the data should direct their requests to these organizations.
The functions for fidelity evaluations utilized in this study have been made publicly available as a Python package. The source code is accessible on GitHub at https://github.com/SCAI-BIO/syndat. The code for privacy evaluations as well as for each synthetization model are available publicly in the respective cited repositories.

https://zenodo.org/records/13929256

## Acknowledgements

This work was done as part of the NFDI4Health Consortium (www.nfdi4health.de). We gratefully acknowledge the financial support of the Deutsche Forschungsgemeinschaft (DFG, German Research Foundation) - project number 442326535.

Data collection and sharing for this project was funded by the Alzheimer’s Disease Neuroimaging Initiative (ADNI) (National Institutes of Health Grant U01 AG024904) and DOD ADNI (Department of Defense award number W81XWH-12-2-0012). ADNI is funded by the National Institute on Aging, the National Institute of Biomedical Imaging and Bioengineering, and through generous contributions from the following: AbbVie, Alzheimer’s Association; Alzheimer’s Drug Discovery Foundation; Araclon Biotech; BioClinica, Inc.; Biogen; Bristol-Myers Squibb Company; CereSpir, Inc.; Cogstate; Eisai Inc.; Elan Pharmaceuticals, Inc.; Eli Lilly and Company; EuroImmun; F. Hoffmann-La Roche Ltd and its affiliated company Genentech, Inc.; Fujirebio; GE Healthcare; IXICO Ltd.; Janssen Alzheimer Immunotherapy Research & Development, LLC.; Johnson & Johnson Pharmaceutical Research & Development LLC.; Lumosity; Lundbeck; Merck & Co., Inc.; Meso Scale Diagnostics, LLC.; NeuroRx Research; Neurotrack Technologies; Novartis Pharmaceuticals Corporation; Pfizer Inc.; Piramal Imaging; Servier; Takeda Pharmaceutical Company; and Transition Therapeutics. The Canadian Institutes of Health Research is providing funds to support ADNI clinical sites in Canada. Private sector contributions are facilitated by the Foundation for the National Institutes of Health (www.fnih.org). The grantee organization is the Northern California Institute for Research and Education, and the study is coordinated by the Alzheimer’s Therapeutic Research Institute at the University of Southern California. ADNI data are disseminated by the Laboratory for Neuro Imaging at the University of Southern California.

## Author Contributions

HF and CB designed the project. HGN, KO and JR generated the synthetic data. TA implemented methods for fidelity and utility evaluation. KO and FP conceptualized the approach to privacy evaluation and its integration with the overall project design. KO implemented methods for privacy evaluation. TA, KO, US, FP and HF interpreted the results. TA, CB, KO, HGN, JR, AFN, US, FP and HF contributed to the content and revision of the manuscript.

## Conflict of Interests

The authors declare no competing interests.

## Resource availability

### Lead contact

Requests for further information and resources should be directed to and will be fulfilled by the lead contact, Tim Adams.

### Materials Availability

This study did not generate new unique reagents.

### Data and Code Availability

The data supporting the findings of this study are available from the respective data holding organizations (c.f. section 2.1). Researchers interested in accessing the data should direct their requests to these organizations.

The functions for fidelity evaluations utilized in this study have been made publicly available as a Python package. The source code is accessible on GitHub at https://github.com/SCAI-BIO/syndat.

The code for privacy evaluations as well as for each synthetization model are available publicly in the respective cited repositories.

## Footnotes

1 https://github.com/SCAI-BIO/syndat

2 https://syndat.scai.fraunhofer.de/

3 https://github.com/spring-epfl/synthetic_data_release

## Notes

### Competing Interest Statement

The authors have declared no competing interest.

### Author Declarations

The used datasets are either directly openly available or available upon request at the respective data holding organizations. Access to the Alzheimer's Disease Neuroimaging Initiative (ADNI) can be requested using the IDA portal: Alzheimer's Disease Neuroimaging Initiative (ADNI) The Center for Cancer Registry Data (ZfKD) can make the validated dataset available to third parties upon request, in accordance with paragraph 5 (3) of the Federal Cancer Registry Data Act (BKRG), provided that the applicant demonstrates a legitimate interest, particularly for scientific purposes: https://www.da-ra.de/dara/study/web_show?res_id=626806&mdlang=en&detail=true The Texas Hospital Inpatient Discharge Data Public Use Data File (TEXAS) can be downloaded for public use on their homepage: https://www.dshs.texas.gov/texas-health-care-information-collection/health-data-researcher-information/texas-inpatient-public-use

### Summary of Updates

- The title and abstract were changed - We extended the introduction with an improved literature review - We elaborated on implementation details in the methods section - We included anonymized data in the utility analysis

## References

[1] S. G. Mueller et al., “Ways toward an early diagnosis in Alzheimer’s disease: the Alzheimer’s Disease Neuroimaging Initiative (ADNI),” Alzheimers Dement. J. Alzheimers Assoc., vol. 1, no. 1, pp. 55–66, Jul. 2005, doi: 10.1016/j.jalz.2005.06.003.

[2] “The Cancer Genome Atlas Pan-Cancer analysis project | Nature Genetics.” Accessed: Dec. 20, 2023. [Online]. Available: https://www.nature.com/articles/ng.2764.

[3] M. W. Weiner et al., “Impact of the Alzheimer’s Disease Neuroimaging Initiative, 2004 to 2014,” Alzheimers Dement., vol. 11, no. 7, pp. 865–884, 2015, doi: 10.1016/j.jalz.2015.04.005.

[4] D. P. Veitch et al., “Using the Alzheimer’s Disease Neuroimaging Initiative to improve early detection, diagnosis, and treatment of Alzheimer’s disease,” Alzheimers Dement., vol. 18, no. 4, pp. 824–857, 2022, doi: 10.1002/alz.12422.

[5] K. Helbing, S. Y. Demiroglu, F. Rakebrandt, K. Pommerening, O. Rienhoff, and U. Sax, “A Data Protection Scheme for Medical Research Networks,” Methods Inf. Med., vol. 49, no. 6, pp. 601–607, 2010, doi: 10.3414/ME09-02-0058.

[6] Y. Zhao, M. Li, L. Lai, N. Suda, D. Civin, and V. Chandra, “Federated Learning with Non-IID Data,” 2018, doi: 10.48550/arXiv.1806.00582.

[7] Q. Li, Y. Diao, Q. Chen, and B. He, “Federated Learning on Non-IID Data Silos: An Experimental Study,” in 2022 IEEE 38th International Conference on Data Engineering (ICDE), May 2022, pp. 965–978. doi: 10.1109/ICDE53745.2022.00077.

[8] J. Walonoski et al., “Synthea: An approach, method, and software mechanism for generating synthetic patients and the synthetic electronic health care record,” J. Am. Med. Inform. Assoc. JAMIA, vol. 25, no. 3, pp. 230–238, Mar. 2018, doi: 10.1093/jamia/ocx079.

[9] M. Hernandez, G. Epelde, A. Alberdi, R. Cilla, and D. Rankin, “Synthetic data generation for tabular health records: A systematic review,” Neurocomputing, vol. 493, pp. 28–45, Jul. 2022, doi: 10.1016/j.neucom.2022.04.053.

[10] J. Nalepa, M. Marcinkiewicz, and M. Kawulok, “Data Augmentation for Brain-Tumor Segmentation: A Review,” Front. Comput. Neurosci., vol. 13, p. 83, Dec. 2019, doi: 10.3389/fncom.2019.00083.

[11] J.-F. Rajotte, R. Bergen, D. L. Buckeridge, K. El Emam, R. Ng, and E. Strome, “Synthetic data as an enabler for machine learning applications in medicine,” iScience, vol. 25, no. 11, p. 105331, Oct. 2022, doi: 10.1016/j.isci.2022.105331.

[12] A. B. Levine et al., “Synthesis of diagnostic quality cancer pathology images by generative adversarial networks,” J. Pathol., vol. 252, no. 2, pp. 178–188, Oct. 2020, doi: 10.1002/path.5509.

[13] H. Chhoa et al., “Improvement of an External Predictive Model Based on New Information Using a Synthetic Data Approach,” Neurol. Genet., vol. 9, no. 5, p. e200091, Oct. 2023, doi: 10.1212/NXG.0000000000200091.

[14] T. Gu, J. M. G. Taylor, W. Cheng, and B. Mukherjee, “Synthetic data method to incorporate external information into a current study,” Can. J. Stat., vol. 47, no. 4, pp. 580–603, 2019, doi: 10.1002/cjs.11513.

[15] B. Jelić, R. Grbić, M. Vranješ, and D. Mijić, “Can We Replace Real-World With Synthetic Data in Deep Learning-Based ADAS Algorithm Development?,” IEEE Consum. Electron. Mag., vol. 12, no. 5, pp. 32–38, Sep. 2023, doi: 10.1109/MCE.2021.3083206.

[16] A. D. Nicholson, D. E. Peplow, J. M. Ghawaly, M. J. Willis, and D. E. Archer, “Generation of Synthetic Data for a Radiation Detection Algorithm Competition,” IEEE Trans. Nucl. Sci., vol. 67, no. 8, pp. 1968–1975, Aug. 2020, doi: 10.1109/TNS.2020.3001754.

[17] T. Stadler, B. Oprisanu, and C. Troncoso, “Synthetic Data – Anonymisation Groundhog Day”.

[18] M. Giomi, F. Boenisch, C. Wehmeyer, and B. Tasnádi, “A Unified Framework for Quantifying Privacy Risk in Synthetic Data,” Proc. Priv. Enhancing Technol., vol. 2023, no. 2, pp. 312–328, Apr. 2023, doi: 10.56553/popets-2023-0055.

[19] Z. Zhang, C. Yan, and B. A. Malin, “Membership inference attacks against synthetic health data,” J. Biomed. Inform., vol. 125, p. 103977, Jan. 2022, doi: 10.1016/j.jbi.2021.103977.

[20] D. Chen, N. Yu, Y. Zhang, and M. Fritz, “GAN-Leaks: A Taxonomy of Membership Inference Attacks against Generative Models,” in Proceedings of the 2020 ACM SIGSAC Conference on Computer and Communications Security, Oct. 2020, pp. 343–362. doi: 10.1145/3372297.3417238.

[21] R. J. Chen, M. Y. Lu, T. Y. Chen, D. F. K. Williamson, and F. Mahmood, “Synthetic data in machine learning for medicine and healthcare,” *Nat*. Biomed. Eng., vol. 5, no. 6, Art. no. 6, Jun. 2021, doi: 10.1038/s41551-021-00751-8.

[22] J. M. Abowd and L. Vilhuber, “How Protective Are Synthetic Data?,” in *Privacy in Statistical Databases*, J. Domingo-Ferrer and Y. Saygın, Eds., in Lecture Notes in Computer Science. Berlin, Heidelberg: Springer, 2008, pp. 239–246. doi: 10.1007/978-3-540-87471-3_20.

[23] C. Dwork, “Differential Privacy,” in *Automata, Languages and Programming*, M. Bugliesi, B. Preneel, V. Sassone, and I. Wegener, Eds., in Lecture Notes in Computer Science. Berlin, Heidelberg: Springer, 2006, pp. 1–12. doi: 10.1007/11787006_1.

[24] M. Pereira, M. Kshirsagar, S. Mukherjee, R. Dodhia, J. L. Ferres, and R. de Sousa, “Assessment of differentially private synthetic data for utility and fairness in end-to-end machine learning pipelines for tabular data,” PLOS ONE, vol. 19, no. 2, p. e0297271, Feb. 2024, doi: 10.1371/journal.pone.0297271.

[25] L. Rosenblatt, X. Liu, S. Pouyanfar, E. de Leon, A. Desai, and J. Allen, “Differentially Private Synthetic Data: Applied Evaluations and Enhancements,” Nov. 10, 2020, arXiv: arXiv:2011.05537. Accessed: May 23, 2024. [Online]. Available: http://arxiv.org/abs/2011.05537

[26] T. Li and N. Li, “On the tradeoff between privacy and utility in data publishing,” in Proceedings of the 15th ACM SIGKDD international conference on Knowledge discovery and data mining, Paris France: ACM, Jun. 2009, pp. 517–526. doi: 10.1145/1557019.1557079.

[27] D. Brunelli, S. Kurapati, and L. Gilli, “SURE: A New Privacy and Utility Assessment Library for Synthetic Data,” in 2024 IEEE International Conference on Blockchain (Blockchain), Aug. 2024, pp. 643–648. doi: 10.1109/Blockchain62396.2024.00094.

[28] A. D. Lautrup, T. Hyrup, A. Zimek, and P. Schneider-Kamp, “Syntheval: a framework for detailed utility and privacy evaluation of tabular synthetic data,” Data Min. Knowl. Discov., vol. 39, no. 1, p. 6, Dec. 2024, doi: 10.1007/s10618-024-01081-4.

[29] F. J. Sarmin, A. R. Sarkar, Y. Wang, and N. Mohammed, “Synthetic Data: Revisiting the Privacy-Utility Trade-off,” Jul. 09, 2024, arXiv: arXiv:2407.07926. doi: 10.48550/arXiv.2407.07926.

[30] “Texas Inpatient Public Use Data File (PUDF) | Texas DSHS.” Accessed: Dec. 18, 2023. [Online]. Available: https://www.dshs.texas.gov/texas-health-care-information-collection/health-data-researcher-information/texas-inpatient-public-use

[31] Q. Razi, S. Datta, V. Hassija, G. Chalapathi, and B. Sikdar, “Privacy Utility Tradeoff Between PETs: Differential Privacy and Synthetic Data,” IEEE Trans. Comput. Soc. Syst., pp. 1–12, 2024, doi: 10.1109/TCSS.2024.3479317.

[32] Q. Chen, J. Yang, M. Huang, and Q. Zhou, “ACT-GAN: Radio map construction based on generative adversarial networks with ACT blocks,” IET Commun., vol. 18, no. 19, pp. 1541–1550, 2024, doi: 10.1049/cmu2.12846.

[33] “Using the ADAP Learning Algorithm to Forecast the Onset of Diabetes Mellitus - PMC.” Accessed: Jan. 27, 2025. [Online]. Available: https://pmc.ncbi.nlm.nih.gov/articles/PMC2245318/

[34] L. Gootjes-Dreesbach, M. Sood, A. Sahay, M. Hofmann-Apitius, and H. Fröhlich, “Variational Autoencoder Modular Bayesian Networks for Simulation of Heterogeneous Clinical Study Data,” *Front*. Big Data, vol. 3, 2020, Accessed: Jan. 08, 2024. [Online]. Available: https://www.frontiersin.org/articles/10.3389/fdata.2020.00016

[35] S. Moazemi et al., “NFDI4Health Workflow and Service for Synthetic Data Generation, Assessment and Risk Management,” in *Studies in Health Technology and Informatics*, R. Röhrig, N. Grabe, U. H. Hübner, K. Jung, U. Sax, C. O. Schmidt, M. Sedlmayr, and A. Zapf, Eds., IOS Press, 2024. doi: 10.3233/SHTI240834.

[36] Zentrum Für Krebsregisterdaten (ZfKD) Im Robert Koch-Institut, “Datensatz des ZfKD auf Basis der epidemiologischen Landeskrebsregisterdaten Epi2016_2, verfügbare Diagnosejahre bis 2014.” ZfKD – German Center for Cancer Registry Data at the RKI, 2017. doi: 10.18444/5.03.01.0005.0012.0002.

[37] L. Efremov, S. F. Abera, A. Bedir, D. Vordermark, and D. Medenwald, “Patterns of glioblastoma treatment and survival over a 16-years period: pooled data from the German Cancer Registries,” J. Cancer Res. Clin. Oncol., vol. 147, no. 11, pp. 3381–3390, Nov. 2021, doi: 10.1007/s00432-021-03596-5.

[38] H. Ping, J. Stoyanovich, and B. Howe, “DataSynthesizer: Privacy-Preserving Synthetic Datasets,” in Proceedings of the 29th International Conference on Scientific and Statistical Database Management, in SSDBM ’17. New York, NY, USA: Association for Computing Machinery, Jun. 2017, pp. 1–5. 10.1145/3085504.3091117.

[39] “DataResponsibly/DataSynthesizer.” Accessed: Jun. 10, 2024. [Online]. Available: https://github.com/DataResponsibly/DataSynthesizer

[40] J. Jordon, J. Yoon, and M. van der Schaar, “PATE-GAN: Generating Synthetic Data with Differential Privacy Guarantees,” presented at the International Conference on Learning Representations, Sep. 2018. Accessed: Jan. 08, 2024. [Online]. Available: https://openreview.net/forum?id=S1zk9iRqF7

[41] N. Papernot, S. Song, I. Mironov, A. Raghunathan, K. Talwar, and Ú. Erlingsson, “Scalable Private Learning with PATE,” Feb. 24, 2018, arXiv: arXiv:1802.08908. doi: 10.48550/arXiv.1802.08908.

[42] L. Kühnel, et al., “Synthetic data generation for a longitudinal cohort study -- Evaluation, method extension and reproduction of published data analysis results,” May 12, 2023, *arXiv*: arXiv:2305.07685.doi: 10.48550/arXiv.2305.07685.

[43] D. Heckerman, “Bayesian Networks for Data Mining,” Data Min. Knowl. Discov., vol. 1, no. 1, pp. 79–119, Mar. 1997, doi: 10.1023/A:1009730122752.

[44] A. Nazábal, P. M. Olmos, Z. Ghahramani, and I. Valera, “Handling incomplete heterogeneous data using VAEs,” Pattern Recognit., vol. 107, p. 107501, Nov. 2020, doi: 10.1016/j.patcog.2020.107501.

[45] “Synthetic Data – Anonymisation Groundhog Day | USENIX.” Accessed: May 16, 2024. [Online]. Available: https://www.usenix.org/conference/usenixsecurity22/presentation/stadler

[46] L. Sweeney, “k-ANONYMITY: A MODEL FOR PROTECTING PRIVACY,” Int. J. Uncertain. Fuzziness Knowl.-Based Syst., May 2012, doi: 10.1142/S0218488502001648.

[47] M. Abadi et al., “Deep Learning with Differential Privacy,” in Proceedings of the 2016 ACM SIGSAC Conference on Computer and Communications Security, in CCS ’16. New York, NY, USA: Association for Computing Machinery, Oct. 2016, pp. 308–318. doi: 10.1145/2976749.2978318.

[48] F. K. Dankar and K. El Emam, “The application of differential privacy to health data,” in Proceedings of the 2012 Joint EDBT/ICDT Workshops, Berlin Germany: ACM, Mar. 2012, pp. 158–166. doi: 10.1145/2320765.2320816.

[49] C. Dwork, N. Kohli, and D. Mulligan, “Differential Privacy in Practice: Expose your Epsilons!,” J. Priv. Confidentiality, vol. 9, no. 2, Art. no. 2, Oct. 2019, doi: 10.29012/jpc.689.

[50] C. M. Bowen and J. Snoke, “Comparative Study of Differentially Private Synthetic Data Algorithms from the NIST PSCR Differential Privacy Synthetic Data Challenge,” Oct. 12, 2020, arXiv: arXiv:1911.12704. Accessed: Jun. 06, 2024. [Online]. Available: http://arxiv.org/abs/1911.12704

[51] D. McClure and J. P. Reiter, “Differential Privacy and Statistical Disclo-sure Risk Measures: An Investigation with Binary Synthetic Data,” 2012.

[52] D. Freedman and P. Diaconis, “On the histogram as a density estimator:L 2 theory,” Z. Für Wahrscheinlichkeitstheorie Verwandte Geb., vol. 57, no. 4, pp. 453–476, Dec. 1981, doi: 10.1007/BF01025868.

[53] F. E. Harrell Jr, R. M. Califf, D. B. Pryor, K. L. Lee, and R. A. Rosati, “Evaluating the Yield of Medical Tests,” JAMA, vol. 247, no. 18, pp. 2543–2546, May 1982, doi: 10.1001/jama.1982.03320430047030.

[54] R. Shokri, M. Stronati, C. Song, and V. Shmatikov, “Membership Inference Attacks against Machine Learning Models,” Mar. 31, 2017, *arXiv*: arXiv:1610.05820. doi: 10.48550/arXiv.1610.05820.

[55] “Article 29 working party ‘Opinion 05/2014 on Anonymisation Techniques’.” [Online]. Available: https://ec.europa.eu/justice/article-29/documentation/opinion-recommendation/files/2014/wp216_en.pdf

[56] “Privacy Risk in Machine Learning: Analyzing the Connection to Overfitting | IEEE Conference Publication | IEEE Xplore.” Accessed: May 17, 2024. [Online]. Available: https://ieeexplore.ieee.org/document/8429311

[57] M. Halilovic, T. Meurers, K. Otte, and F. Prasser, “Are You the Outlier? Identifying Targets for Privacy Attacks on Health Datasets,” in Digital Health and Informatics Innovations for Sustainable Health Care Systems, IOS Press, 2024, pp. 1224–1225. doi: 10.3233/SHTI240631.

[58] statice/anonymeter. (Jun. 05, 2024). Python. Statice. Accessed: Jun. 12, 2024. [Online]. Available: https://github.com/statice/anonymeter

[59] S. M. Lundberg and S.-I. Lee, “A Unified Approach to Interpreting Model Predictions”.

[60] R. McKenna, B. Mullins, D. Sheldon, and G. Miklau, “AIM: An Adaptive and Iterative Mechanism for Differentially Private Synthetic Data,” Jun. 13, 2024, arXiv: arXiv:2201.12677. doi: 10.48550/arXiv.2201.12677.

[61] “Towards formalizing the GDPR’s notion of singling out | PNAS.” Accessed: Jun. 27, 2024. [Online]. Available: https://www.pnas.org/doi/full/10.1073/pnas.1914598117

[62] N. Hollmann, S. Müller, K. Eggensperger, and F. Hutter, “TabPFN: A Transformer That Solves Small Tabular Classification Problems in a Second,” arXiv.org. Accessed: Oct. 08, 2024. [Online]. Available: https://arxiv.org/abs/2207.01848v6

[63] B. Kaabachi et al., “A Scoping Review of Privacy and Utility Metrics in Medical Synthetic Data,” Aug. 28, 2024, *medRxiv*. doi: 10.1101/2023.11.28.23299124.

