## Supplementary Material for "On the Fidelity versus Privacy and Utility Trade-Off of Synthetic Patient Data"

### VAMBN implementation details

VAMBN, in addition to general hyper-parameter configuration, incorporates domain knowledge by organizing variables into modular groupings during the Encoding/Decoding process. These groupings allow for more precise modeling of variable relationships. Furthermore, constraints are applied through blacklists and whitelists to control the edges in the Bayesian network, ensuring that domain-relevant relationships are enforced while excluding non-meaningful connections. This framework allows for a tailored representation of complex data, capturing key dependencies.

For easier reproducibility, we provide configuration files for all three datasets in a supplementary Zenodo repository[1]. Configuration files can be used in the 1.0.0 public VAMBN version[2] to reproduce the results of this study.

For further details on how to apply these configurations, please refer to the documentation of the corresponding github repository.

### Supplementary Figures and Tables

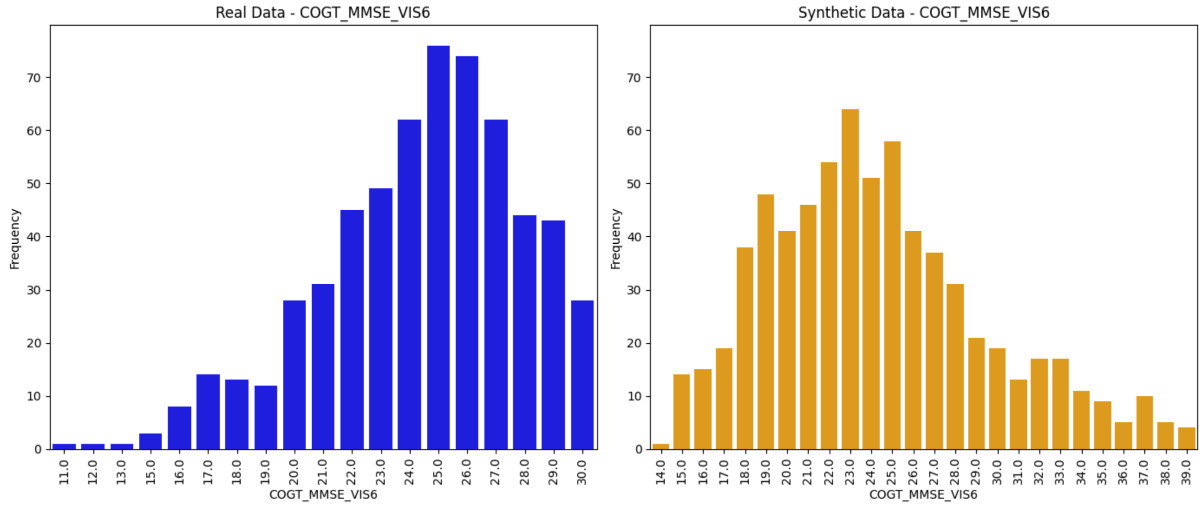


**Figure S1**: Distribution of real and synthetic values for MMSE test.

---

[1] https://zenodo.org/records/13929256
[2] https://github.com/nfdi4health/docker-vambn/releases/tag/v1.0.0

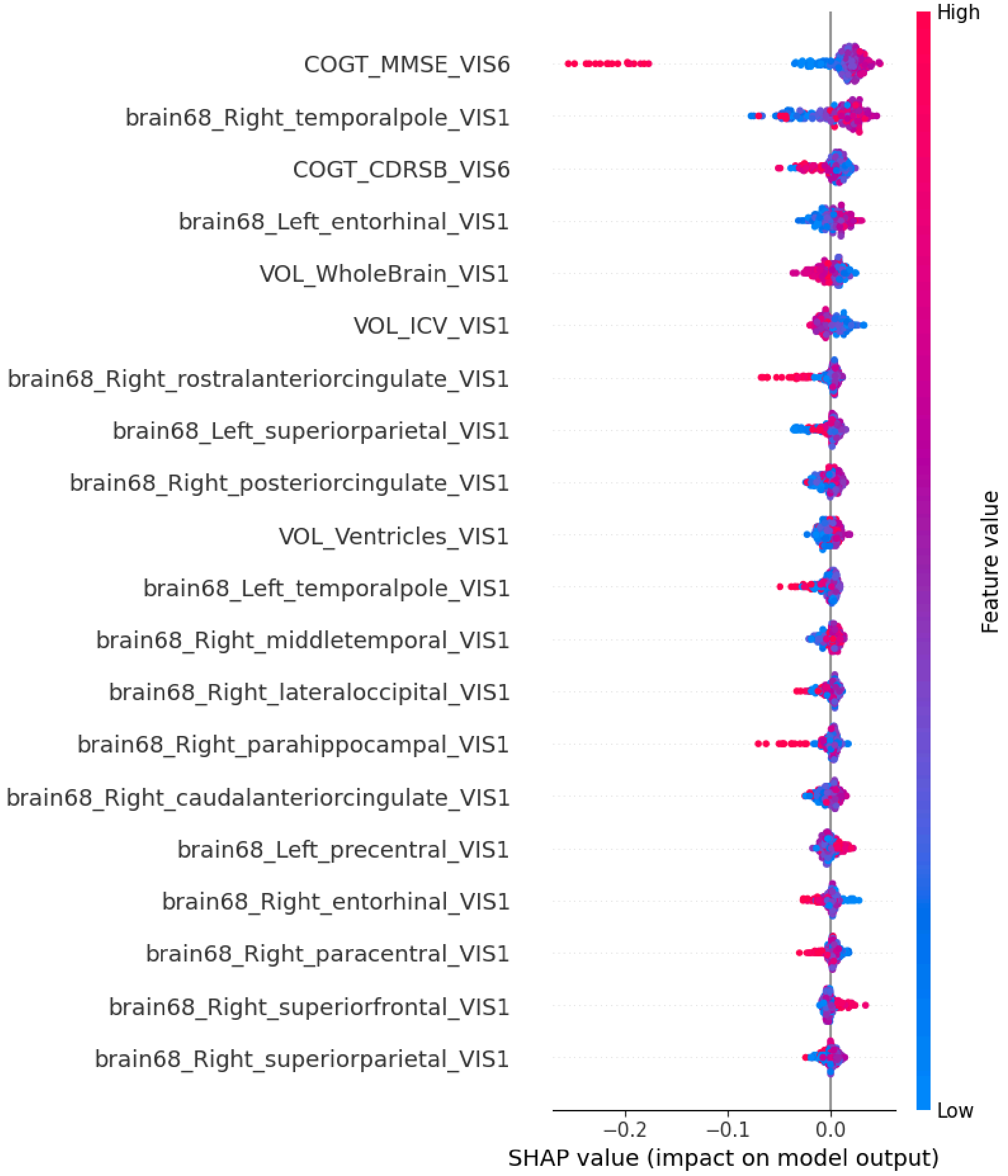


**Figure S2**: SHAP Analysis of RF model discriminating between real and synthetic data for the ADNI dataset.

| | sdims | ydims | nbatch | lrates | wdecay | loss | epochs | epsilon budget |
|---|---|---|---|---|---|---|---|---|
| **Texas VAMBN** | 2 | 2 | 16 | 0.0021 | 0.0026 | -12.2388 | 349 | - |
| **VAMBN DP low** | 2 | 2 | 16 | 0.0021 | 0.0026 | -12.2388 | 349 | 5 |
| **VAMBN DP high** | 2 | 2 | 16 | 0.0021 | 0.0026 | -12.2388 | 349 | 0.45 |
| **RKI VAMBN** | 6 | 7 | 128 | 0.01 | 0.01 | -20.1422 | 116 | - |
| **VAMBN DP low** | 6 | 7 | 128 | 0.01 | 0.01 | -20.1422 | 116 | 5 |
| **VAMBN DP high** | 6 | 7 | 128 | 0.01 | 0.01 | -20.1422 | 116 | 1.02 |

**Table S1:** VAMBN parameterizations.

| | epsilon | delta | degree | n_bins | n_iters | nbatch | lrates |
|---|---|---|---|---|---|---|---|
| **Bayesian Net** | - | - | - | 25 | - | - | - |
| **PateGAN** | 1.0 | 1e-5 | - | - | 100 | 128 | 1e-4 |
| **PrivBayes** | 1.0 | - | 1 | 10 | - | - | - |
| **Sanatizer NHS** | - | - | - | 10 | - | - | - |

**Table S2:** Other parameterizations.

| Dataset | Quasi Identifiers |
|---|---|
| **ADNI** | SA_PTEDUCAT_VIS1, SA_PTGENDER_VIS1, SA_PTETHCAT_VIS1, SA_PTRACCAT_VIS1, SA_PTMARRY_VIS1, SA_AGE_VIS1 |
| **RKI** | TOD, SEX |
| **TEXAS** | PAT_STATE, SEX_CODE, RACE, ETHNICITY, PAT_AGE |

**Table S3:** Quasi-Identifiers used for by SanatizerNHS for each dataset

| Target Model | Privacy Gain Average Records | Privacy Gain Outlier Records | |
|---|---|---|---|
| | mean (CI) | mean (CI) | Abs. Diff |
| **BayesianNet** | 1.00 (0.67, 1.33) | 1.07 (0.71, 1.42) | -0.07 |
| **PateGan (ε=1.0)** | - | - | - |
| **PrivBayes (ε=1.0)** | 1.00 (1.00, 1.00) | 1.00 (1.00, 1.00) | 0.0 |
| **SanitiserNHS (k=10)** | - | - | - |
| **VAMBN** | 1.13 (0.76, 1.51) | 1.20 (0.92, 1.48) | -0.07 |
| **VAMBN DP (ε=1.2)** | 0.87 (0.54, 1.19) | 1.13 (0.81, 1.46) | -0.03 |
| **VAMBN DP (ε=5.0)** | 0.87 (0.44, 1.29) | 1.07 (0.84, 1.30) | -0.2 |

**Table S4:** Privacy Gain for average and outlier targets for the generated synthetic ADNI data.

| Target Model | Privacy Gain Average Records | Privacy Gain Outlier Records | |
|---|---|---|---|
| | mean (CI) | mean (CI) | Abs. Diff |
| **BayesianNet** | 1.03 (0.95, 1.10) | 0.67 (0.55, 0.76) | 0.36 |
| **PateGan (ε=1.0)** | 1.04 (0.89, 1.25) | 0.92 (0.77, 1.18) | 0.13 |
| **PrivBayes (ε=1.0)** | 1.01 (0.82, 1.20) | 1.14 (0.97, 1.31) | -0.13 |
| **SanitiserNHS (k=10)** | 0.99 (0.93, 1.02) | 0.86 (0.77, 0.97) | 0.13 |
| **VAMBN** | 1.04 (0.91, 1.17) | 0.83 (0.68, 0.96) | 0.21 |
| **VAMBN DP (ε=0.45)** | 1.01 (0.91, 1.12) | 0.95 (0.85, 1.07) | 0.06 |
| **VAMBN DP (ε=5.0)** | 1.04 (1.01, 1.28) | 0.81 (0.74, 0.98) | 0.23 |

**Table S5:** Privacy Gain for average and outlier targets for the generated synthetic TEXAS data.

| | Privacy Gain Average Records | Privacy Gain Outlier Records | |
|---|---|---|---|
| **Target Model** | **mean (CI)** | **mean (CI)** | **Abs. Diff** |
| **BayesianNet** | 1.09 (0.91, 1.28) | 0.99 (0.84, 1.14) | 0.10 |
| **PateGan (ε=1.0)** | 0.91 (0.74, 1.08) | 0.98 (0.81, 1.14) | -0.07 |
| **PrivBayes (ε=1.0)** | 0.86 (0.70, 1.03) | 1.01 (0.83, 1.19) | -0.15 |
| **SanitiserNHS (k=10)** | 0.91 (0.83, 1.00) | 0.98 (0.93, 1.02) | -0.07 |
| **VAMBN** | 1.00 (0.92, 1.08) | 0.94 (0.85, 1.02) | 0.06 |
| **VAMBN DP (ε=0.45)** | 1.03 (0.88, 1.18) | 0.93 (0.81, 1.05) | 0.09 |
| **VAMBN DP (ε=5.0)** | 0.99 (0.87, 1.11) | 1.04 (0.88, 1.20) | -0.05 |

**Table S6:** Privacy Gain for average and outlier targets for the generated synthetic RKI data.

| | # Cat | # Num | Target Variable | Omitted Variables |
|---|---|---|---|---|
| **ADNI** | 5 | 79 | **COGT_MMSE_VIS1**<br>(<24) | - all vars after baseline visit<br>- (***_VIS6**, *_**VIS12**, '*_**VIS24**)<br>- all vars with a missingness >90%<br>- Diagnose (**SA_DX_VIS1**)<br>**- SA_APOE4_VIS1**<br>**- SA_PTEDUCAT_VIS1**<br>**- SA_PTGENDER_VIS1**<br>**- SA_PTETHCAT_VIS1**<br>**- SA_PTRACCAT_VIS1**<br>**- SA_PTMARRY_VIS1**<br>**- SA_AGE_VIS1**<br>**- SA_APOE4_VIS1**<br>**- A_FDG_VIS1** |
| **RKI** | 7 | 2 | **DAYS_TILL_DEATH_OR_END** | none |
| **TEXAS** | 11 | 7 | **ILLNESS_SEVERITY, RISK_MORTALITY** | none |

**Table S7:** Further variable information for the predictions tasks.
